# A Deep Learning-Derived Insulin Resistance Index for Cardiovascular Risk Prediction: A Prospective Cohort Study with External Validation in Chinese and US Populations

**DOI:** 10.64898/2026.08.10.26360145

**Authors:** Yaqian Mao, Jing Lin, Ao Zhou, Shuhang Zeng, Dang Yang, Wei Lin, Junping Wen, Wenjie Yang, Gang Chen

**Author notes:** These authors contributed equally to this work (Yaqian Mao, Jing Lin). Correspondence: Gang Chen, Wenjie Yang.

## Abstract

**Background:** Existing insulin resistance (IR) indices are predominantly developed in diabetic cohorts, limiting their generalizability. We developed a novel deep neural network–derived IR index (DNN-IR) using a Mixture-of-Experts (MoE) framework and evaluated its predictive performance for incident cardiovascular disease (CVD) and mortality in general populations.

**Methods:** We utilized data from three cohorts: the cross-sectional REACTION study (Fujian subcohort, 2011-2012) for DNN-IR derivation and internal validation; and two prospective cohorts, NHANES (1999-2018, linked to the National Death Index) and CHARLS (2011-2018), for external validation. The DNN-IR was developed using a deep learning model based on a Mixture-of-Experts (MoE) architecture, trained on the REACTION dataset. We evaluated the DNN-IR’s utility in predicting incident CVD, cardiovascular mortality, and non-cardiovascular mortality among 13,889 NHANES and 7,047 CHARLS participants. Predictive performance was assessed via the area under the receiver operating characteristic curve (AUC). Multivariable logistic regression, restricted cubic splines, and Kaplan-Meier analyses characterized the associations between DNN-IR and clinical outcomes.

**Results:** In the REACTION cohort, DNN-IR demonstrated superior predictive performance for atherosclerotic outcomes, achieving AUROCs of 0.89 (training) and 0.84 (internal validation). In the external CHARLS cohort (median follow-up: 7 years; 1,135 incident CVD cases [16.1%]), DNN-IR yielded AUROCs of 0.72 for incident CVD and 0.77 for all-cause mortality. Fully adjusted models showed that each 1-SD increment in DNN-IR was associated with a 23% higher CVD risk (OR=1.23, 95% CI: 1.14-1.32), exhibiting a predominantly linear dose-response relationship (*P*-nonlinearity=0.453). In NHANES, DNN-IR robustly predicted cardiovascular (AUROC=0.77) and all-cause mortality (AUROC=0.72), alongside specific mortalities like diabetes (0.91), Alzheimer’s disease (0.88), and kidney disease (0.96). Higher DNN-IR levels correlated with stepwise increases in cumulative mortality (log-rank *P*<0.001).

**Conclusions:** The MoE-derived DNN-IR index demonstrated robust and stable performance in predicting atherosclerosis, incident CVD, cardiovascular mortality, and all-cause mortality in the general population. Further validation in larger, more diverse cohorts is warranted to support its broad clinical applicability.

## Introduction

Cardiovascular diseases (CVDs), primarily comprising ischemic heart disease and stroke, represent the leading cause of death and disability worldwide^[1]^. According to the 2023 Global Burden of Disease (GBD) study, CVDs have remained the leading cause of death and disability worldwide for 33 consecutive years, with the burden continuing to escalate. Globally, the number of people living with CVD surged from 311 million in 1990 to 626 million in 2023—an increase of over 100%^[2]^. Concurrently, annual deaths rose from 13.1 million to 19.2 million, equivalent to approximately 52,600 lives lost daily to CVD^[2]^. A CVD surveillance study based on the Chinese population indicates a heavy burden of CVD incidence in China, with the crude incidence rate estimated at 620.33 per 100,000 population in 2023^[3]^. As the world’s most populous country, China accounts for the largest absolute number of CVD deaths globally, and the disease burden continues to grow increasingly heavy^[4]^. Therefore, early identification and intervention targeting CVD risk factors hold significant importance for reducing the global disease burden.

Atherosclerosis represents the core pathological mechanism underlying the increasing incidence of CVD events^[5,6]^. A growing body of research indicates that arterial stiffness also serves as an independent predictor of CVD onset and mortality, directly impacting the cardiovascular system^[7,8]^. Brachial-ankle pulse wave velocity (baPWV), a extensively validated non-invasive clinical metric, effectively assesses arterial stiffness and has been demonstrated in epidemiological studies to correlate closely with CVD events and all-cause mortality^[9]^. Given the long-standing and insidious nature of its pathogenesis, there is an urgent need to develop simple and reliable biomarkers for early diagnosis and intervention.

Insulin resistance (IR) is closely associated with the onset and progression of cardiovascular diseases and mortality ^[10–12]^. To date, the hyperinsulinemic-euglycemic clamp remains the gold standard for assessing insulin resistance; however, its cumbersome procedure and high cost limit its widespread clinical application^[13]^. In clinical practice, the Homeostatic Model Assessment (HOMA-IR) is a commonly used method, calculated primarily from fasting insulin and glucose levels ^[14]^. Since HOMA-IR relies on insulin measurements, its broad clinical adoption faces challenges. In recent years, an increasing number of insulin resistance surrogate markers have emerged, such as the triglyceride-glucose (TyG) index and its derivatives, demonstrating strong predictive power and stability^[15,16]^. A study by Zhang et al. explored the associations between different IR indices and all-cause and cardiovascular mortality, finding that the TyG index was significantly associated with both outcomes^[17]^. Another study by Hu et al. showed that the triglyceride-glucose-body mass index (TyG-BMI) and the TyG index were the two optimal predictors of obesity, and that a stacked machine learning model incorporating TyG-BMI effectively predicted stroke risk^[18]^. Additionally, research by Li et al. indicated that individuals with higher TyG indices had a significantly increased risk of arterial stiffness ^[19]^.

Although various IR surrogate indices currently exist and demonstrate good predictive performance, their relationships with arterial stiffness, incident cardiovascular events, and cardiovascular mortality remain unclear. Moreover, most existing studies have focused solely on the impact of a single IR index on outcomes, without comparing the performance across different indices. The majority of current research consists of single-center, small-sample cross-sectional studies, lacking the ability to establish causality and often missing external validation across diverse populations.

With the rapid advancement of artificial intelligence, machine learning (ML) has demonstrated powerful predictive capabilities and shown promising performance across various diseases. Several ML algorithms are now employed in clinical research, including Logistic Regression (LR), Random Forest (RF), Extreme Gradient Boosting (XGBoost), Light Gradient Boosting Machine (LightGBM), and Adaptive Boosting (AdaBoost). However, the comparative predictive performance among these algorithms requires further analysis. This study aims to develop a novel insulin resistance predictor using neural networks and compare its predictive performance against existing IR indices. Beyond evaluating the performance of this deep neural network-derived insulin resistance index (DNN-IR) in predicting arterial stiffness, incident cardiovascular events, and cardiovascular mortality, we also aim to assess its predictive value for other chronic diseases.

## Methods

### Selection and description of participants

The study dataset comprised one cross-sectional dataset and two prospective cohort datasets. The cross-sectional data were derived from the Fujian subcohort of the Risk Evaluation of cAncers in Chinese diabeTic Individuals: a lONgitudinal study (REACTION). Participants in the two prospective national cohorts were drawn from the China Health and Retirement Longitudinal Study (CHARLS) in China and the National Health and Nutrition Examination Survey (NHANES) in the United States. In the first phase of this study, the REACTION dataset was randomly split into a training set (approximately 80%, *n* = 5,000) and an internal test set (20%, *n* = 1,298). The training set was utilized for the development and selection of the DNN-IR index. The NHANES and CHARLS datasets were used for external validation to evaluate the clinical utility of the DNN-IR index in predicting incident CVD events, cardiovascular mortality, all-cause mortality, and mortality from other diseases. The study design is illustrated in **Figure 1**.

**Fig. 1.**
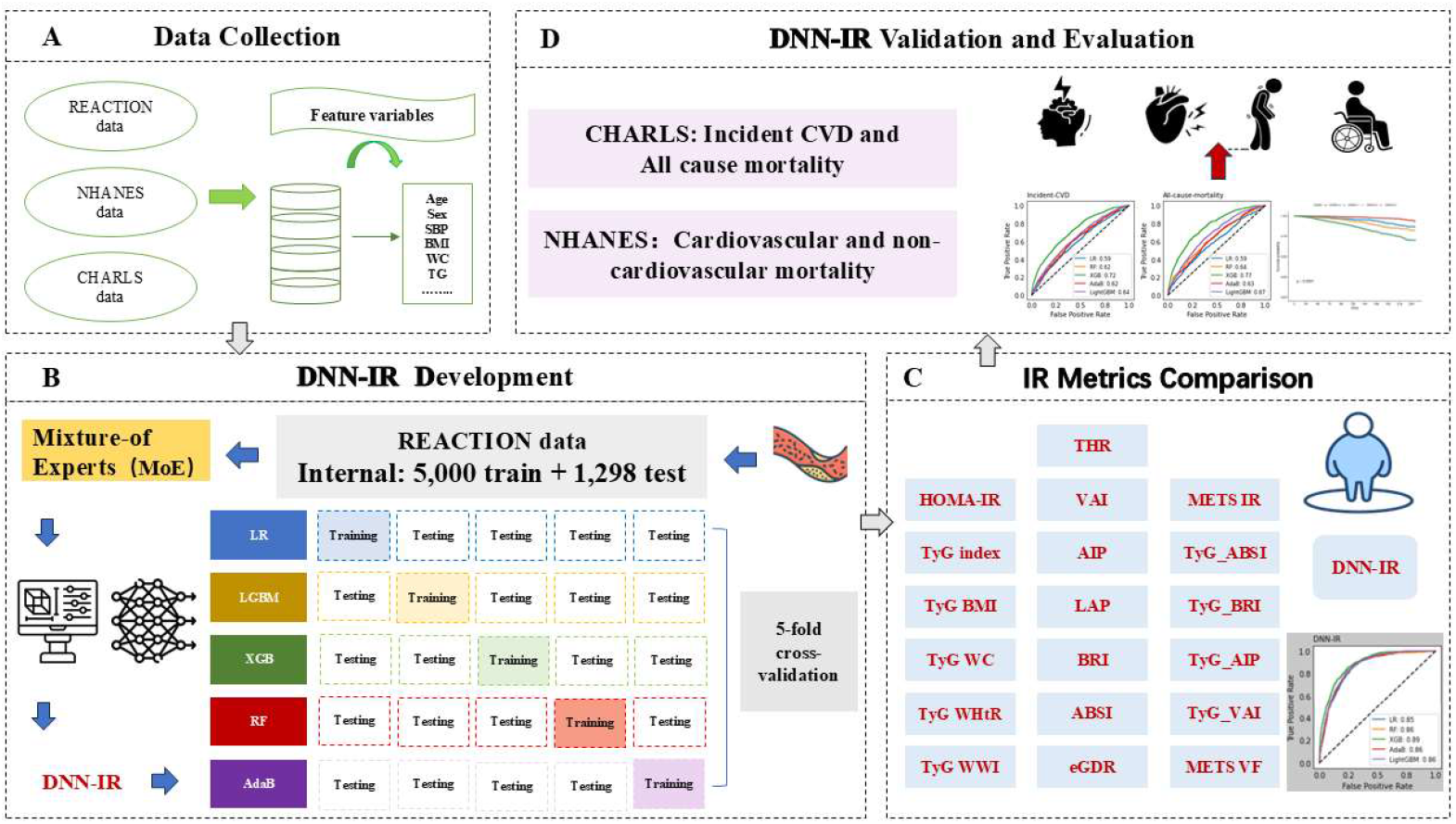
Study Design and Workflow. **Note:** (A) Overview of data sources from the REACTION, NHANES, and CHARLS cohorts. (B) Development of the DNN-IR using a Mixture-of-Experts (MoE) deep learning model trained on the REACTION dataset. (C) Performance comparison between the DNN-IR and 19 established traditional IR indices within the REACTION cohort. (D) External validation of the DNN-IR for predicting incident CVD and all-cause mortality in the independent CHARLS and NHANES cohorts. **Abbreviations:** MoE, Mixture-of-Experts; CVD, Cardiovascular disease; DNN-IR, Deep neural network–derived insulin resistance index; IR, Insulin resistance; CHARLS, China Health and Retirement Longitudinal Study; NHANES, National Health and Nutrition Examination Survey; REACTION, Risk Evaluation of cAncers in Chinese diabeTic Individuals: a lONgitudinal study; HOMA-IR, Homeostatic model assessment for insulin resistance; THR, Triglyceride to high-density lipoprotein cholesterol ratio; METS-IR, Metabolic score for insulin resistance; VAI, Visceral adiposity index; AIP, Atherogenic index of plasma; LAP, Lipid accumulation product; BRI, Body roundness index; ABSI, A body shape index; eGDR, Estimated glucose disposal rate; TyG, Triglyceride and glucose index; TyG-BMI, TyG combined with body mass index; TyG-WC, TyG combined with waist circumference; TyG-WHtR, TyG combined with waist-to-height ratio; TyG-WWI, TyG combined with weight-adjusted waist index; TyG-BRI, TyG combined with body roundness index; TyG-ABSI, TyG combined with a body shape index; TyG-AIP, TyG combined with atherogenic index of plasma; TyG-VAI, TyG combined with visceral adiposity index; METS-VF, Metabolic score for visceral fat.

The Fujian subcohort of the REACTION study was conducted from January 2011 to January 2012, initially enrolling 6,437 participants who completed brachial-ankle pulse wave velocity (baPWV) measurements. Participants were excluded if they met any of the following criteria: (1) age < 35 years; (2) missing data on laboratory tests, anthropometric measurements, or demographic characteristics; or (3) extreme physiological values, defined as a waist-to-height ratio (WHtR) < 0.3 or > 0.8, or a body mass index (BMI) < 15 or > 55 kg/m². Ultimately, 6,298 participants were included.

The CHARLS is a longitudinal cohort study conducted from 2011 to 2018, comprising four survey waves (2011, 2013, 2015, and 2018). Initially, 11,847 participants who provided blood samples during the first wave were enrolled. Exclusion criteria included: (1) age < 35 years; (2) missing anthropometric or demographic data at baseline (wave 1), or the presence of extreme values (WHtR < 0.3 or > 0.8; BMI < 15 or > 55 kg/m²); (3) prevalent CVD at baseline (wave 1); (4) lack of follow-up records in waves 2, 3, or 4; (5) missing demographic, anthropometric, or laboratory data during follow-up; or (6) missing data on incident CVD or vital status. A total of 7,047 participants were ultimately included in the study.

The NHANES is a continuous, nationally representative cross-sectional survey. We collected complete NHANES datasets from 1999 to 2020 and constructed a longitudinal follow-up cohort by linking mortality data via the National Death Index (NDI) through the National Center for Health Statistics (NCHS). Follow-up was censored on December 31, 2019. Initially, 59,064 participants who underwent NHANES examinations between 1999 and 2020 were included. Exclusion criteria were: (1) age < 35 years; (2) missing data on laboratory tests, anthropometric measurements, or demographic characteristics; (3) missing vital status data; or (4) extreme values (BMI < 15 or > 55 kg/m²; WHtR < 0.3 or > 0.8). Finally, 13,889 participants were included. The detailed inclusion and exclusion criteria for the three cohorts are illustrated in **Figure 2**.

**Fig. 2.**
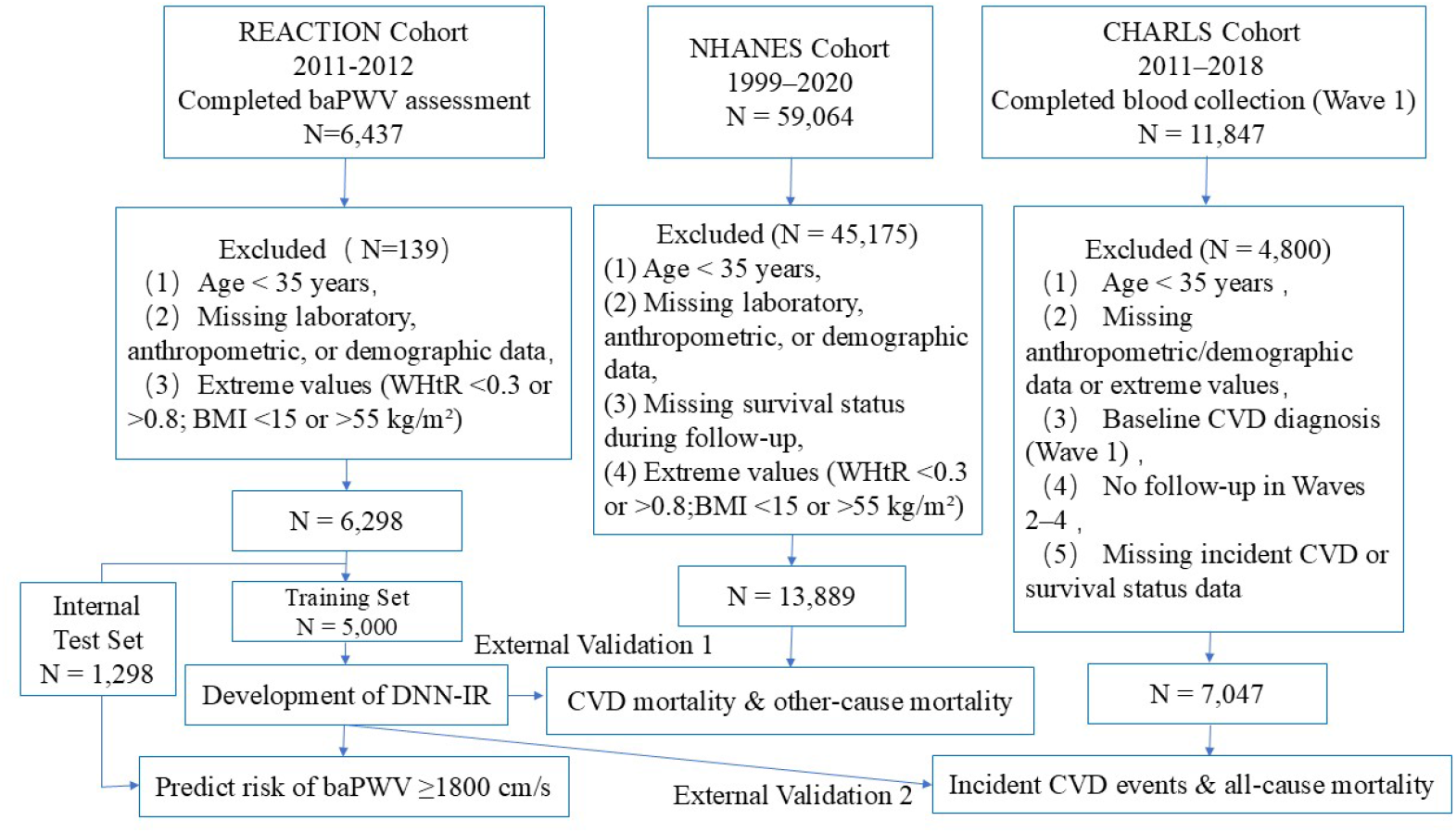
Flowchart of the study population. **Note:** Three independent cohorts were utilized: the REACTION cohort (2011–2012) for the development and internal validation of the DNN-IR index to predict arterial stiffness (baPWV ≥1800 cm/s); the CHARLS cohort (2011–2018) for the first external validation (predicting incident CVD and all-cause mortality); and the NHANES cohort (1999–2020) for the second external validation (predicting CVD and cause-specific mortality). Detailed exclusion criteria and final sample sizes are provided for each cohort. **Abbreviations**: baPWV, Brachial-ankle pulse wave velocity; CVD, Cardiovascular disease; DNN-IR, Deep neural network–derived insulin resistance index; CHARLS, China Health and Retirement Longitudinal Study; NHANES, National Health and Nutrition Examination Survey; REACTION, Risk Evaluation of cAncers in Chinese diabeTic Individuals: a lONgitudinal study.

### Data collection and measurements

Data collection and processing were standardized across the REACTION, CHARLS, and NHANES cohorts to ensure consistency. Based on previous literature and expert consensus, we selected 22 covariates known to significantly influence arterial stiffness and cardiovascular disease risk. These included demographic characteristics (age, sex, education level, marital status), lifestyle factors (smoking and drinking status), anthropometric measurements (weight, height, waist circumference [WC], BMI, WHtR), and clinical/laboratory variables (systolic blood pressure [SBP], diastolic blood pressure [DBP], hypertension, diabetes, dyslipidemia, fasting plasma glucose [FPG], HbA1c, triglycerides [TG], high-density lipoprotein cholesterol [HDL-C], low-density lipoprotein cholesterol [LDL-C], and total cholesterol [TC]). Education was categorized into three levels (below high school, high school, and college or above); marital status was dichotomized (married vs. unmarried); and smoking and alcohol consumption were classified into three categories (never, former, and current).

Anthropometric measurements were obtained following standard protocols. WC was measured at the end of normal expiration while participants stood upright with feet slightly apart. Brachial-ankle pulse wave velocity (baPWV) was measured bilaterally and simultaneously using an automated waveform analyzer (Omron Colin BP-203PRE III; Omron Healthcare, Kyoto, Japan). The mean value of the left and right baPWV was used for statistical analyses. Arterial stiffness was defined as a baPWV ≥ 1800 cm/s, according to established diagnostic criteria^[20]^.

Based on previous studies, we selected 19 established IR indices, including HOMA-IR, METS-IR, VAI, LAP, BRI, TyG, and various TyG-derived composite indices (e.g., TyG-BMI, TyG-WC, TyG-WHtR). Detailed definitions, calculation formulas, and references for all 19 indices are provided in **Supplementary Table S1**.

### Definitions of insulin resistance indices

To automatically extract a highly predictive and clinically accessible surrogate IR index from routine clinical variables, we developed a dual-branch deep learning feature generation framework using the REACTION dataset. Nine clinical features—FPG, TG, HDL-C, weight, WC, height, history of hypertension, HbA1c, and sex—were included as inputs. These were derived from the constituent variables of the 19 traditional IR indices. Notably, fasting insulin was excluded as it is not routinely measured in real-world clinical practice. Using baPWV as the reference standard label for arterial stiffness, the dataset was randomly split into a training set (*n* = 5,000) and an internal test set (*n* = 1,298) at a 7:3 ratio.

Subsequently, the newly developed DNN-IR was compared with the 19 traditional IR indices. Five machine learning algorithms—LR, RF, XGBoost, LightGBM, and AdaBoost—were employed to evaluate the predictive performance of each IR index for arterial stiffness. Model performance was assessed using the area under the receiver operating characteristic curve (AUC) on the internal test set. The primary objective was to identify the optimal IR index with the highest predictive accuracy.

### Definition of clinical outcomes

The primary outcome was incident CVD, including myocardial infarction, hemorrhagic stroke, and ischemic stroke, while secondary outcomes comprised cardiovascular, all-cause, and other cause-specific mortalities. In the CHARLS cohort, incident CVD and mortality were ascertained via self-reports through 2018; incident CVD was defined by a physician’s diagnosis or medical treatment for heart disease or stroke (with the diagnosis date as the event date), and mortality was based on self-reported vital status and date of death. In the NHANES cohort, vital status and underlying causes of death were determined via linkage to the National Death Index through December 31, 2019, and classified using *International Classification of Diseases, Tenth Revision* (ICD-10) codes. Cardiovascular mortality included heart diseases (I00–I09, I11, I13, I20–I51) and cerebrovascular diseases (I60–I69). Other cause-specific mortalities were defined by codes for diabetes mellitus (E10–E14), Alzheimer’s disease (G30), chronic lower respiratory diseases (J40–J47), malignant neoplasms (C00–C97), unintentional injuries (V01–X59, Y85–Y86), influenza and pneumonia (J09–J18), and nephritis, nephrotic syndrome, and nephrosis (N00–N07, N17–N19, N25–N27).

### Ethics approval and consent to participate

The original REACTION, CHARLS, and NHANES studies were approved by their respective ethics committees, and all participants provided written informed consent. Because the present study utilized de-identified, publicly available data from the CHARLS and NHANES databases without direct participant intervention, the requirement for additional informed consent was waived. The investigators strictly adhered to the data use agreements of both databases, ensuring full compliance with ethical standards and data security requirements.

### Statistical Analysis

Participants in the REACTION cohort were stratified into quartiles of baPWV, while those in the CHARLS and NHANES cohorts were dichotomized by incident CVD and CVD mortality, respectively. Baseline characteristics were expressed as mean ± standard deviation (SD) or median (interquartile range [IQR]) for continuous variables, and frequencies (percentages) for categorical variables. Group differences were compared using one-way ANOVA or Student’s *t*-test for normally distributed data, the Kruskal-Wallis or Mann-Whitney *U* test for skewed data, and the Pearson chi-square test for categorical variables.

Multivariable logistic regression was employed to evaluate the association between DNN-IR and incident CVD, reporting odds ratios (ORs) with 95% confidence intervals (CIs). Three sequential models were constructed: Model 1 adjusted for demographics (sex, age, marital status, education); Model 2 further adjusted for lifestyle and hemodynamic factors (smoking, alcohol consumption, SBP, DBP); and Model 3 additionally adjusted for the lipid profile (TG, TC, HDL-C, LDL-C). To avoid mathematical coupling and overadjustment bias, we strictly excluded the constituent components of DNN-IR (FPG, weight, WC, hypertension, HbA1c), their derived indices (BMI, WHtR), and potential mediators (diabetes, dyslipidemia) from the covariates. Multicollinearity was assessed using the adjusted generalized variance inflation factor (GVIF)^[21]^. All VIF values were <5 (**Supplementary Table S2**). Although TC (GVIF = 3.95) and LDL-C (GVIF = 3.61) showed moderate collinearity, both were retained given their clinical significance. To ensure collinearity did not substantially bias the effect estimates, we constructed two alternative models: Model S1 (excluding TC) and Model S2 (excluding LDL-C). A change in OR exceeding 10% would indicate substantial bias.

Restricted cubic spline (RCS) logistic regression with 4 knots (5th, 35th, 65th, and 95th percentiles) was used to explore the dose-response relationship between DNN-IR and incident CVD risk. Subgroup analyses were conducted across 12 stratifications (age, sex, marital status, education, smoking, alcohol, SBP, DBP, LDL-C, HDL-C, TG, and TC) to evaluate potential effect modifications. In the NHANES cohort, Kaplan-Meier survival curves were plotted, and the log-rank test was used to compare survival distributions.

### Construction and selection of the DNN-IR index

The DNN-IR was developed using a deep learning model integrating a sparse mask mechanism and a MoE architecture. The model adaptively weighted input features via a learnable mask matrix and utilized parallel multi-layer perceptron (MLP) "expert" subnetworks to generate 14 candidate IR features. To balance predictive performance and clinical interpretability, L1 regularization was introduced to enforce feature sparsity, and Kullback-Leibler (KL) divergence loss was applied for knowledge distillation, ensuring the new features retained key clinical information (detailed architecture in **Supplementary Table S3**).

Following training, univariable logistic regression was applied to evaluate the association of each candidate feature with baPWV in the training set. The feature yielding the highest AUC and relying solely on 5 routine variables (FPG, weight, WC, hypertension history, and HbA1c) was selected and defined as the DNN-IR. Its performance was internally validated in the test set and externally validated for predicting incident CVD in CHARLS, and cardiovascular and all-cause mortality in NHANES.

All statistical analyses were performed using R (versions 4.3.3) and Python (version 3.10). Two-sided *P* < 0.05 was considered statistically significant.

## Results

### Baseline characteristics of participants

The study population selection process is illustrated in **Figure 2**. In the REACTION cohort, 6,298 participants were included in the final analysis, of whom 3,369 (53.5%) were female, with a mean age of 52.81 ± 8.96 years. Baseline characteristics stratified by baPWV quartiles are presented in **Table 1**. Compared with the lowest quartile (Q1), participants in the higher baPWV quartiles (Q2–Q4) had lower education levels, a higher proportion of unmarried individuals, elevated blood pressure, greater BMI, higher levels of TG, TC, LDL-C, FPG, and HbA1c, lower HDL-C levels, and higher prevalences of hypertension, dyslipidemia, and diabetes (all *P* < 0.001). Additionally, all 19 traditional IR indices and the DNN-IR exhibited a progressive increase across baPWV quartiles, indicating that greater arterial stiffness was associated with higher IR. Baseline characteristics of the CHARLS cohort stratified by incident CVD status are summarized in **Supplementary Table S4**, and those of the NHANES cohort stratified by CVD mortality status are presented in **Supplementary Table S5**.

**Table 1.** Baseline characteristics of individuals classifed by quartiles of the baPWV in REACTION study.

| Characteristics | Overall | Quartiles of the BaPWV |  |  |  | P-value |
| --- | --- | --- | --- | --- | --- | --- |
|  |  | Quartile 1 | Quartile 2 | Quartile 3 | Quartile 4 |  |
| n | 6298 | 1576 | 1574 | 1573 | 1575 |  |
| Gender |  |  |  |  |  | <0.001 |
| Male | 2929 (46.5) | 596 (37.8) | 820 (52.1) | 794 (50.5) | 719 (45.7) |  |
| Female | 3369 (53.5) | 980 (62.2) | 754 (47.9) | 779 (49.5) | 856 (54.3) |  |
| Age, years | 52.81 (8.96) | 47.274 (6.095) | 50.194 (7.297) | 53.854 (8.273) | 59.933 (8.540) | <0.001 |
| Education level |  |  |  |  |  | <0.001 |
| Less than high school | 3918 (62.2) | 831 (52.728) | 942 (59.848) | 999 (63.509) | 1146 (72.762) |  |
| High school | 1539 (24.4) | 454 (28.807) | 383 (24.333) | 395 (25.111) | 307 (19.492) |  |
| Some college or higher | 841 (13.4) | 291 (18.464) | 249 (15.820) | 179 (11.380) | 122 ( 7.746) |  |
| Current married |  |  |  |  |  | <0.001 |
| No | 352 ( 5.6) | 65 ( 4.124) | 68 ( 4.320) | 91 ( 5.785) | 128 ( 8.127) |  |
| Yes | 5946 (94.4) | 1511 (95.876) | 1506 (95.680) | 1482 (94.215) | 1447 (91.873) |  |
| Smoking status |  |  |  |  |  | <0.001 |
| Never | 4225 (67.1) | 1140 (72.335) | 978 (62.135) | 1017 (64.654) | 1090 (69.206) |  |
| Former | 364 ( 5.8) | 56 ( 3.553) | 94 ( 5.972) | 93 ( 5.912) | 121 ( 7.683) |  |
| Current | 1709 (27.1) | 380 (24.112) | 502 (31.893) | 463 (29.434) | 364 (23.111) |  |
| Drinking status |  |  |  |  |  | <0.001 |
| Never | 2845 (45.2) | 691 (43.845) | 656 (41.677) | 700 (44.501) | 798 (50.667) |  |
| Former | 358 ( 5.7) | 86 ( 5.457) | 96 ( 6.099) | 83 ( 5.277) | 93 ( 5.905) |  |
| Current | 3095 (49.1) | 799 (50.698) | 822 (52.224) | 790 (50.223) | 684 (43.429) |  |
| SBP, mmHg | 135.19 (19.37) | 119.526(11.327) | 129.337 (12.990) | 138.117 (14.851) | 153.786 (18.693) | <0.001 |
| DBP, mmHg | 78.99 (10.71) | 72.364 (8.083) | 77.547 (8.763) | 81.008 (9.867) | 85.056 (11.512) | <0.001 |
| Height (cm) | 158.98 (7.75) | 159.827 (7.140) | 160.123 (7.685) | 159.234 (7.789) | 156.749 (7.923) | <0.001 |
| Weight (kg) | 61.48 (10.02) | 60.204 (9.329) | 61.847 (10.055) | 62.737 (10.479) | 61.135 (10.038) | <0.001 |
| BMI, kg/m<sup>2</sup> | 24.27 (3.19) | 23.516 (2.884) | 24.057 (3.077) | 24.683 (3.345) | 24.830 (3.280) | <0.001 |
| WC, cm | 81.12 (8.64) | 78.133 (7.946) | 80.460 (8.547) | 82.350 (8.764) | 83.538 (8.314) | <0.001 |
| WHtR | 0.51 (0.05) | 0.489 (0.047) | 0.503 (0.051) | 0.518 (0.054) | 0.534 (0.054) | <0.001 |
| Diabetes | 998 (15.8) | 85 ( 5.393) | 155 ( 9.848) | 290 (18.436) | 468 (29.714) | <0.001 |
| Hypertension | 2710 (43.0) | 116 ( 7.360) | 419 (26.620) | 855 (54.355) | 1320 (83.810) | <0.001 |
| Dyslipidemia | 4090 (64.9) | 818 (51.904) | 1001 (63.596) | 1105 (70.248) | 1166 (74.032) | <0.001 |
| FPG, mmol/L | 5.93 (1.42) | 5.485 (0.814) | 5.758 (1.081) | 6.011 (1.357) | 6.461 (1.971) | <0.001 |
| HbA1c, % | 5.82 (0.96) | 5.525 (0.627) | 5.687 (0.715) | 5.885 (0.950) | 6.164 (1.299) | <0.001 |
| TG, mmol/L | 1.75 (1.34) | 1.419 (1.023) | 1.718 (1.295) | 1.901 (1.541) | 1.981 (1.394) | <0.001 |
| TC, mmol/L | 5.19 (0.98) | 4.977 (0.915) | 5.133 (0.897) | 5.273 (1.017) | 5.359 (1.046) | <0.001 |
| HDL-c, mmol/L | 1.43 (0.33) | 1.460 (0.323) | 1.422 (0.330) | 1.414 (0.322) | 1.415 (0.341) | <0.001 |
| LDL-c, mmol/L | 3.01 (0.80) | 2.857 (0.757) | 2.994 (0.731) | 3.060 (0.814) | 3.119 (0.860) | <0.001 |
| HOMA-IR | 1.75 (1.45) | 1.441 (1.005) | 1.597 (1.334) | 1.868 (1.586) | 2.108 (1.693) | <0.001 |
| TyG-index | 8.82 (0.63) | 8.570 (0.551) | 8.781 (0.606) | 8.903 (0.639) | 9.024 (0.647) | <0.001 |
| THR | 1.38 (1.38) | 1.081 (1.026) | 1.358 (1.360) | 1.516 (1.628) | 1.556 (1.386) | <0.001 |
| METS-IR | 36.00 (6.67) | 33.954 (5.892) | 35.607 (6.493) | 36.943 (6.930) | 37.516 (6.745) | <0.001 |
| VAI | 2.10 (2.06) | 1.663 (1.497) | 2.001 (1.886) | 2.290 (2.452) | 2.446 (2.206) | <0.001 |
| AIP | 0.02 (0.30) | -0.069 (0.279) | 0.016 (0.300) | 0.054 (0.309) | 0.082 (0.295) | <0.001 |
| LAP | 37.82 (38.20) | 26.755 (26.821) | 35.154 (35.490) | 42.707 (44.005) | 46.680 (41.133) | <0.001 |
| BRI | 3.62 (1.07) | 3.200 (0.895) | 3.463 (0.988) | 3.754 (1.081) | 4.071 (1.110) | <0.001 |
| ABSI | 0.08 (0.00) | 0.075 (0.004) | 0.076 (0.004) | 0.077 (0.004) | 0.079 (0.005) | <0.001 |
| eGDR | 9.19 (2.22) | 10.831 (1.347) | 9.876 (1.930) | 8.652 (2.129) | 7.388 (1.722) | <0.001 |
| METS-VF | 6.39 (0.59) | 6.082 (0.575) | 6.305 (0.575) | 6.484 (0.549) | 6.671 (0.500) | <0.001 |
| TyG-BMI | 214.75 (36.64) | 202.077(32.067) | 211.907 (35.166) | 220.432 (37.980) | 224.607 (36.965) | <0.001 |
| TyG-WC | 717.64(108.40) | 671.318(95.033) | 708.603(105.417) | 735.204(109.309) | 755.469(104.721) | <0.001 |
| TyG-WHtR | 4.52 (0.67) | 4.201 (0.565) | 4.426 (0.623) | 4.620 (0.670) | 4.826 (0.669) | <0.001 |
| TyG-WWI | 91.64 (10.07) | 86.575 (8.270) | 90.169 (9.124) | 92.953 (9.736) | 96.861 (10.114) | <0.001 |
| TyG-ABSI | 0.68 (0.07) | 0.647 (0.058) | 0.672 (0.065) | 0.688 (0.067) | 0.710 (0.069) | <0.001 |
| TyG-BRI | 32.19 (10.57) | 27.592 (8.569) | 30.603 (9.573) | 33.640 (10.660) | 36.921 (10.954) | <0.001 |
| TyG-AIP | 0.36 (2.79) | -0.450 (2.462) | 0.306 (2.754) | 0.664 (2.911) | 0.913 (2.809) | <0.001 |
| TyG-VAI | 19.54 (22.01) | 14.920 (15.473) | 18.485 (19.853) | 21.580 (26.611) | 23.197 (23.614) | <0.001 |
| DNN-IR | -0.36 (1.72) | -1.749 (1.019) | -0.948 (1.424) | 0.021 (1.522) | 1.241 (1.215) | <0.001 |

### Performance of the DNN-IR and selection of the optimal IR metric

The DNN-IR was constructed using a dual-branch deep learning framework integrating a sparse mask mechanism and a MoE architecture. This novel IR index comprises five routine clinical variables: FPG, weight, WC, history of hypertension, and HbA1c (detailed variable generation procedures and source codes are available at https://github.com/deepalchemist/dnn-ir). To evaluate its predictive performance for arterial stiffness (baPWV) against existing IR indices, five machine learning algorithms were employed. The DNN-IR demonstrated robust predictive performance in both the training set (**Figure 3**) and the internal test set (**Supplementary Figure 1**). In the training set, the DNN-IR achieved excellent discrimination across all algorithms, with AUC values ranging from 0.85 (LR) to 0.89 (XGBoost). Similarly, in the internal test set, the predictive performance remained consistently high, with AUC values ranging from 0.83 to 0.84 across the five algorithms.

**Fig. 3.**
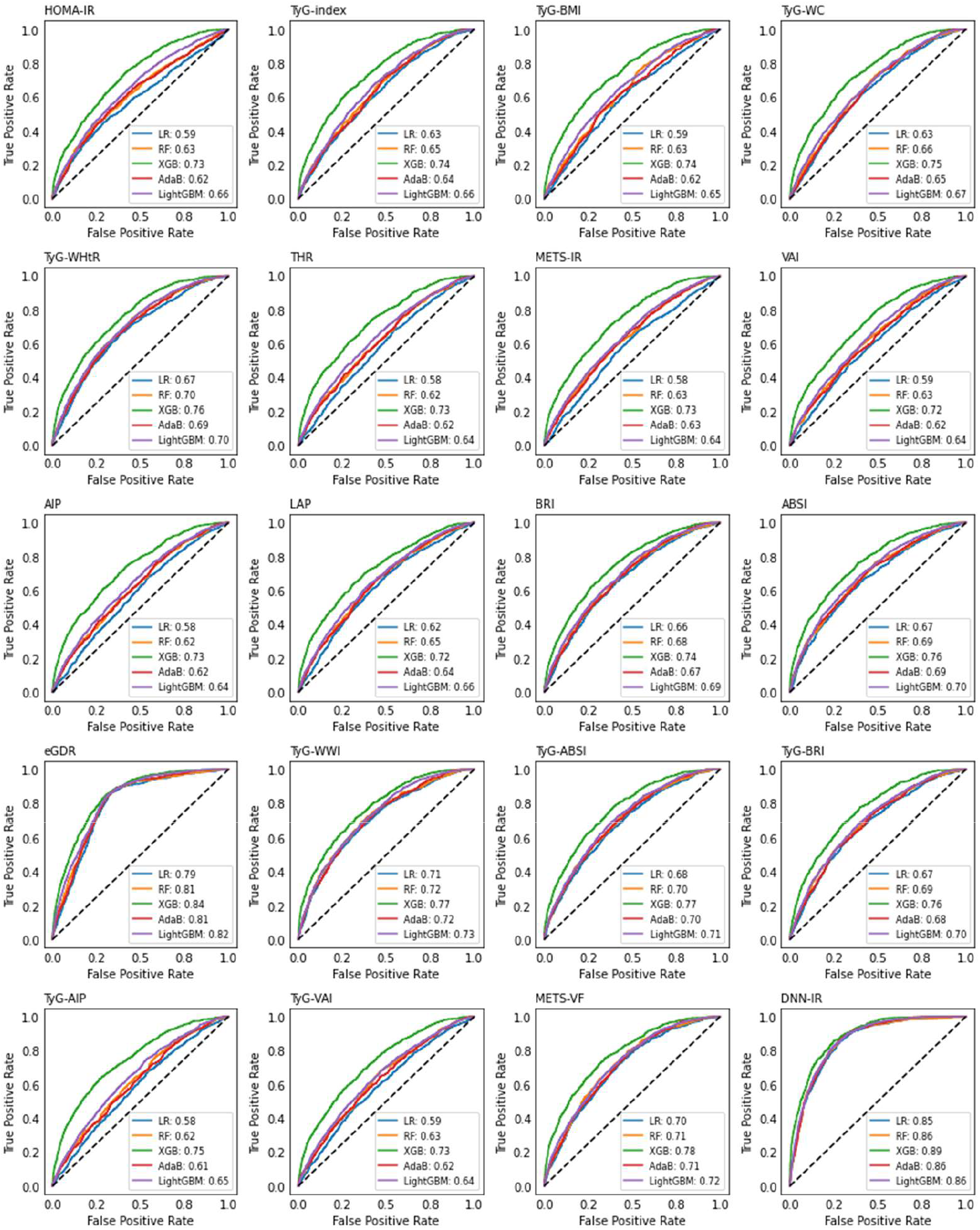
AUC values of base classifiers using DNN-IR and conventional insulin resistance indices for predicting atherosclerosis (baPWV≥1800cm/s) risk in the REACTION training set. LightGBM, Light gradient boosting machine; XGB, Extreme gradient boosting; RF, Random forest; AdaB, Adaptive boosting; LR, Logistic regression; baPWV, Brachial-ankle pulse wave velocity; DNN-IR, Deep neural network-derived insulin resistance index; IR, Insulin resistance; AUC, Area under the receiver operating characteristic curve; REACTION, Risk evaluation of cancers in Chinese diabetic individuals: a longitudinal study; HOMA-IR, Homeostatic model assessment for insulin resistance; THR, Triglyceride to high-density lipoprotein cholesterol ratio; METS-IR, Metabolic score for insulin resistance; VAI, Visceral adiposity index; AIP, Atherogenic index of plasma; LAP, Lipid accumulation product; BRI, Body roundness index; ABSI, A body shape index; eGDR, Estimated glucose disposal rate; TyG, Triglyceride and glucose index; TyG-BMI, TyG combined with body mass index; TyG-WC, TyG combined with waist circumference; TyG-WHtR, TyG combined with waist-to-height ratio; TyG-WWI, TyG combined with weight-adjusted waist index; TyG-BRI, TyG combined with body roundness index; TyG-ABSI, TyG combined with a body shape index; TyG-AIP, TyG combined with atherogenic index of plasma; TyG-VAI, TyG combined with visceral adiposity index; METS-VF, Metabolic score for visceral fat; DNN-IR, Deep neural network-derived insulin resistance index.

In the CHARLS cohort, a total of 7,047 participants were enrolled at baseline. Over a 7-year follow-up period (from wave 1 in 2011 to wave 4 in 2018), 1,135 participants (16.1%) developed incident CVD, including 821 (11.7%) cases of heart disease and 423 (6.0%) cases of stroke, while 611 (8.7%) died (detailed in Supplementary Table 2). Participants with higher DNN-IR levels generally exhibited more pronounced insulin resistance profiles.

The associations between DNN-IR and incident CVD risk are summarized in Table 2. In the unadjusted model, participants in the highest DNN-IR quartile (Q4) had a significantly higher risk of incident CVD compared with those in the lowest quartile (Q1) (OR = 2.32, 95% CI: 1.93–2.80). This positive association remained robust after sequential adjustment for potential confounders. In the fully adjusted model (Model 3), the corresponding ORs (95% CIs) across increasing quartiles were 1.26 (1.03–1.54) for Q2, 1.39 (1.13–1.71) for Q3, and 1.77 (1.43–2.20) for Q4. Significant upward trends across increasing DNN-IR quartiles were observed in all models (*P* for trend < 0.001). Furthermore, each 1-SD increment in DNN-IR was associated with a 36% higher risk of incident CVD in the unadjusted model (OR = 1.36, 95% CI: 1.27–1.44) and a 23% higher risk in the fully adjusted model (OR = 1.23, 95% CI: 1.14–1.32).

**Table 2.**
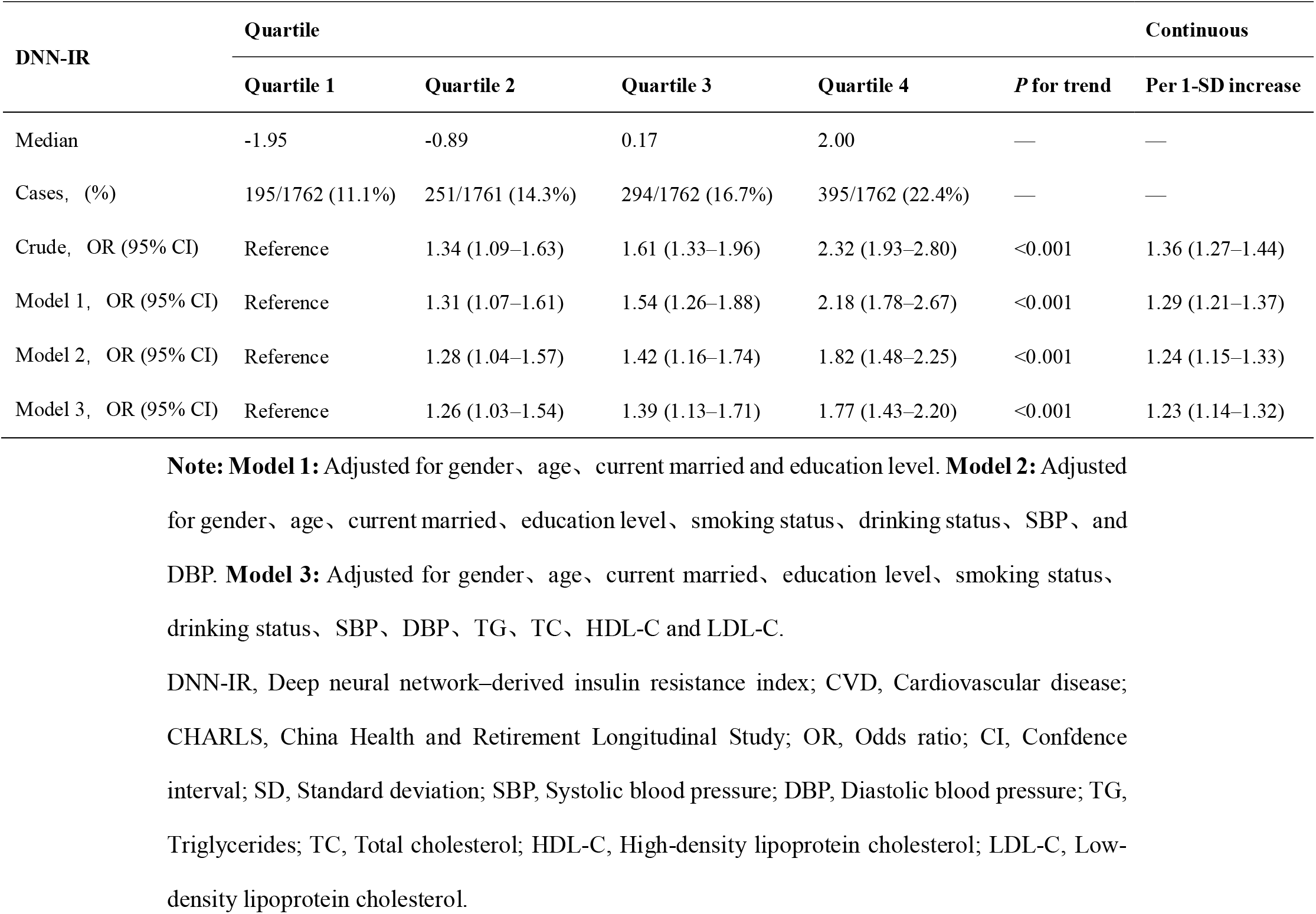
Association between the DNN-IR and incident CVD in the CHARLS cohort.

Sensitivity analyses confirmed the robustness of the DNN-IR–incident CVD association after excluding either TC or LDL-C from the fully adjusted model (**Supplementary Table S6**). The risk of incident CVD for the highest versus lowest DNN-IR quartile remained significantly elevated in both alternative models (excluding TC: OR = 1.76 [95% CI: 1.42–2.19]; excluding LDL-C: OR = 1.76 [95% CI: 1.42– 2.19]; both *P* for trend < 0.001). Compared with the primary model (Q4 vs. Q1: OR = 1.77; per 1-SD increment: OR = 1.23), the effect estimates changed by <0.5%, well below the 10% threshold for substantial bias. These findings indicate that the moderate collinearity between TC and LDL-C did not materially confound the observed associations, supporting the robustness of our primary results.

**Figure 4** illustrates the multivariable-adjusted dose-response relationships between DNN-IR and incident cardiovascular events in the CHARLS cohort. Fully adjusted restricted cubic spline (RCS) analyses revealed a significant, predominantly linear positive association between DNN-IR and total CVD risk (*P*-overall < 0.001, *P*-nonlinearity = 0.453; **Figure 4A**). Similarly, predominantly linear associations were observed for incident heart disease (*P*-overall < 0.001, *P*-nonlinearity = 0.423; **Figure 4B**) and stroke (*P*-overall < 0.001, *P*-nonlinearity = 0.090; **Figure 4C**). The risks of CVD and stroke consistently increased across the entire DNN-IR spectrum. Notably, for incident heart disease, the risk remained relatively stable (OR ≈ 1.0) at lower DNN-IR scores (< −1.0) before increasing sharply beyond the median DNN-IR level.

**Fig. 4.**
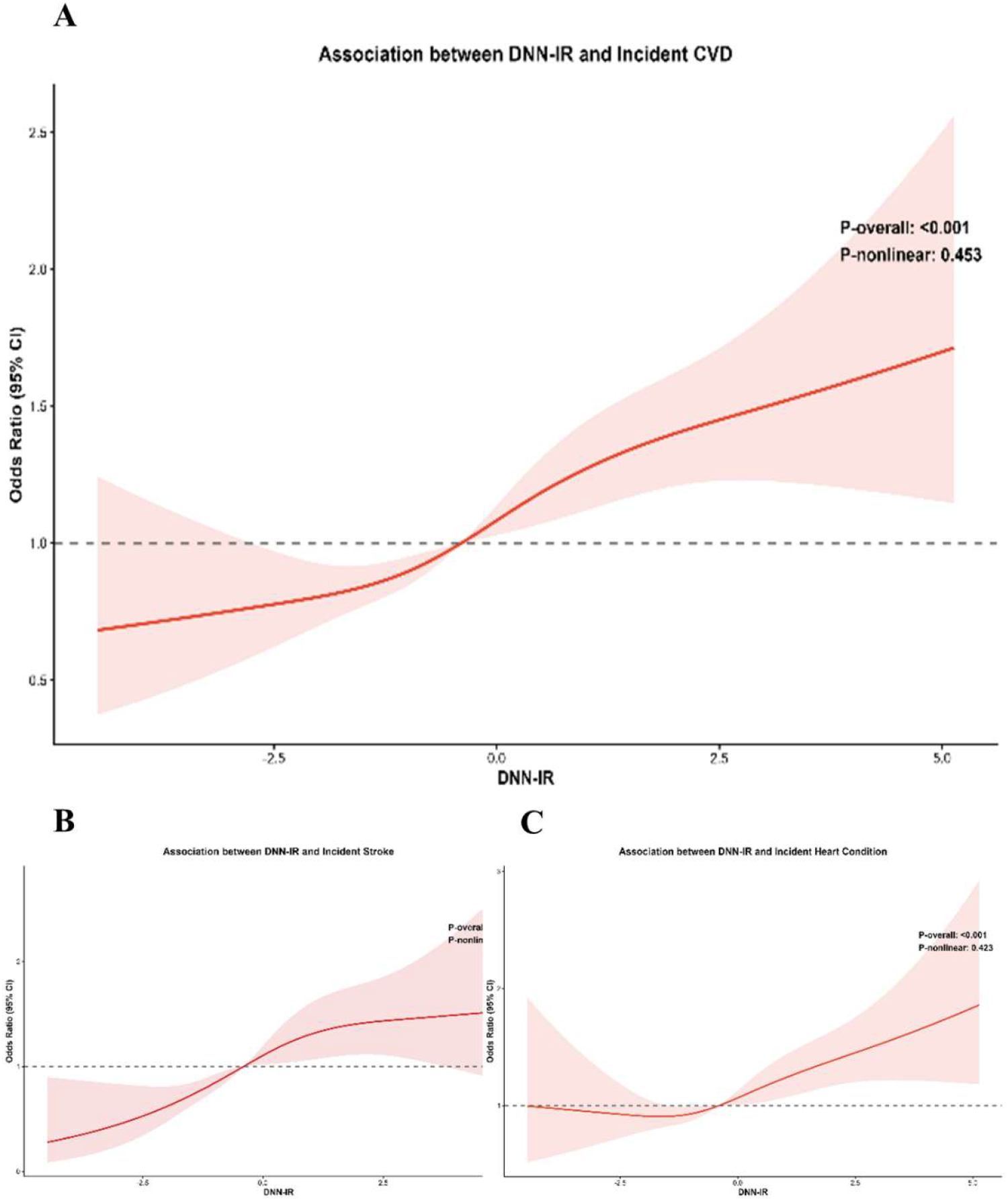
Associations of DNN-IR with incident CVD (A), incident heart disease (B), and incident stroke (C) in the CHARLS cohort. Models were adjusted for sex, age, marital status, education level, smoking status, drinking status, SBP, DBP, TG, TC, HDL-C, and LDL-C. **Abbreviations:** DNN-IR, Deep neural network–derived insulin resistance index; CVD, Cardiovascular disease; CHARLS, China Health and Retirement Longitudinal Study; SBP, Systolic blood pressure; DBP, Diastolic blood pressure; TG, Triglycerides; TC, Total cholesterol; HDL-C, High-density lipoprotein cholesterol; LDL-C, Low-density lipoprotein cholesterol; CI, Confdence interval.

To evaluate the generalizability and robustness of the DNN-IR index, we conducted external validation in the CHARLS cohort (**Supplementary Figure 2**). The DNN-IR demonstrated significant predictive performance for both incident CVD and all-cause mortality. For incident CVD (**Supplementary Figure 2A**), the AUC values across the evaluated models ranged from 0.59 to 0.72, confirming its utility in identifying individuals at high cardiovascular risk. Furthermore, the index exhibited enhanced predictive capability for all-cause mortality (**Supplementary Figure 2B**), with AUC values ranging from 0.59 to 0.77. These findings underscore that the DNN-IR, by integrating metabolic and anthropometric features, serves as a robust predictor for both specific cardiovascular events and overall mortality risk.

### Subgroup analysis

To further elucidate the association between DNN-IR and the risk of CVD, we conducted a series of subgroup analyses. As illustrated in **Supplementary Figure 3**, the association between DNN-IR and incident CVD was not significantly modified by age, sex, marital status, smoking status, alcohol consumption, SBP, DBP, LDL-C, HDL-C, TG, or TC (all *P* for interaction > 0.05), with the exception of educational level.

### Predictive performance of DNN-IR for mortality outcomes

To comprehensively evaluate the predictive performance of the DNN-IR, we conducted receiver operating characteristic (ROC) analyses for all-cause and cause-specific mortality in the NHANES cohort (**Figure 5**). The DNN-IR demonstrated stable discriminative ability for all-cause mortality (AUC: 0.68–0.72) and robust predictive value for cardiovascular mortality (AUC: 0.69–0.77). Notably, the index exhibited strong predictive capability across a broad spectrum of other cause-specific mortalities. It achieved the highest performance in predicting diabetes-related mortality (AUC: 0.77–0.91), followed by Alzheimer’s disease (AUC: 0.69–0.88), nephritis (AUC: 0.66– 0.96), chronic lower respiratory diseases (AUC: 0.66–0.86), other specific causes (AUC: 0.66–0.76), and cancer (AUC: 0.59–0.75). These findings indicate that the DNN-IR serves not only as an effective insulin resistance metric but also as a comprehensive biomarker for forecasting diverse mortality risks.

**Fig. 5.**
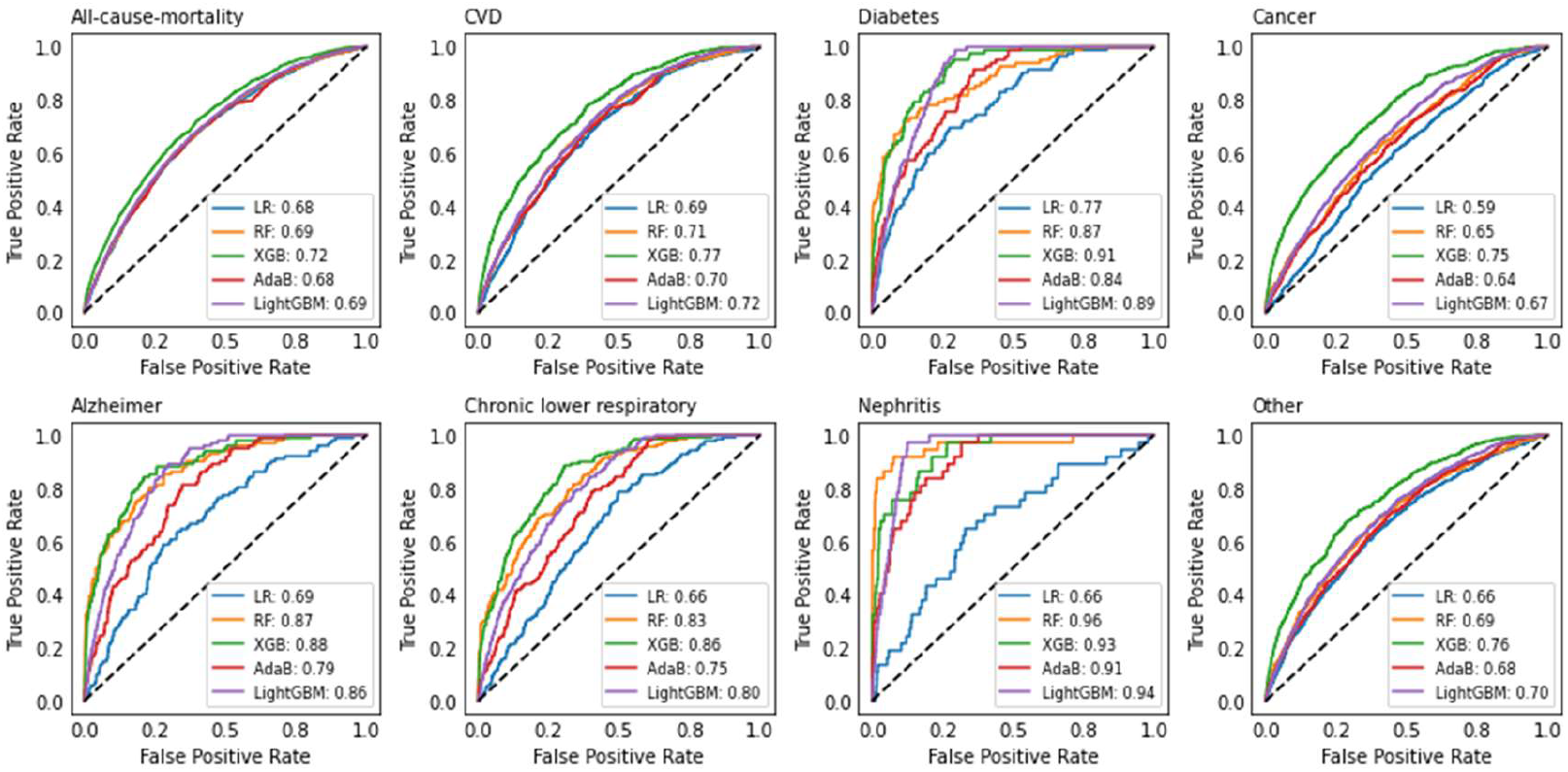
Area under the curve (AUC) values of the base classifier using DNN-IR to predict CVD mortality and mortality from other causes in the NHANES cohort. AUC, Area under the curve; CVD, Cardiovascular disease; DNN-IR, Deep neural network–derived insulin resistance index; NHANES, National Health and Nutrition Examination Survey; AdaB, Adaptive boosting; LightGBM, Light gradient boosting machine; LR, Logistic regression; RF, Random forest; XGB, Extreme gradient boosting.

Participants were stratified into four quartiles (Q1–Q4) based on DNN-IR levels. Supplementary Figure 4A presents the cumulative incidence of CKD-related mortality during follow-up, and Supplementary Figure 4B displays the cumulative incidence of all-cause mortality. The lowest quartile (Q1) served as the reference group. Survival distributions across quartiles were compared using the log-rank test. All P values were < 0.0001, indicating that participants in the highest quartile (Q4) had significantly higher rates of both CKD-related mortality and all-cause mortality compared with the reference group.

## Discussion

In this study, we developed and validated a novel DNN-IR using a sparse mask mechanism integrated with a MoE deep learning framework. The DNN-IR, composed of five readily available clinical variables-FPG, weight, WC, hypertension, and HbA1c — demonstrated superior performance over 19 conventional IR indices in predicting atherosclerosis, as assessed by BaPWV, in a Chinese general population cohort. Furthermore, DNN-IR exhibited robust predictive capacity for incident CVD, cardiovascular mortality, and all-cause mortality in two independent external validation cohorts—the CHARLS and the NHANES. Notably, DNN-IR also showed strong discriminative ability for multiple cause-specific mortalities, including diabetes-related, Alzheimer’s disease, chronic lower respiratory disease, and nephritis-related deaths, underscoring its potential as a comprehensive metabolic health biomarker beyond CVD risk stratification

### DNN-IR and Atherosclerosis: Comparison with Traditional IR Indices

The association between IR and arterial stiffness is well-established^[22][23]^. In the present study, we employed baPWV, a validated non-invasive measure of arterial stiffness with established prognostic value for cardiovascular events^[24][25]^, as the ground-truth label for IR index development. Among the 20 indices evaluated (19 established IR surrogates plus DNN-IR), DNN-IR achieved the highest discriminative performance for atherosclerotic burden (training AUC: 0.89; internal validation AUC: 0.84), followed by the eGDR (training AUC: 0.84; validation AUC: 0.79). This finding is clinically significant because most existing IR indices—including HOMA-IR, TyG index, and their anthropometric derivatives—were originally developed and validated primarily in diabetic populations or for predicting incident diabetes^[26][27]^, and their performance in identifying subclinical atherosclerosis among general populations remains suboptimal. Recent work has similarly demonstrated the superiority of machine learning-derived IR assessments over traditional formula-based indices^[28][29]^.

The superior performance of DNN-IR can be attributed to several factors. First, the sparse mask mechanism enabled automatic feature selection from a broad pool of candidate variables, allowing the model to identify the most informative combination without a priori assumptions about variable importance. The final DNN-IR components — FPG, weight, WC, hypertension, and HbA1c — collectively capture both gluco-metabolic dysregulation (FPG, HbA1c), adiposity (weight, WC), and hemodynamic stress (hypertension), three interconnected pathophysiological domains of insulin resistance. Second, the MoE architecture facilitated the modeling of heterogeneous subpopulations with distinct IR phenotypes, a critical advantage given the well-recognized heterogeneity in IR manifestations across age, sex, and body composition groups.

### DNN-IR and Incident CVD in the CHARLS Cohort

In the CHARLS external validation cohort, DNN-IR demonstrated a robust and independent association with incident CVD over 7 years of follow-up. After full adjustment for demographic, lifestyle, and metabolic confounders, each 1-SD increase in DNN-IR was associated with a 23% higher risk of incident CVD (OR = 1.23, 95% CI: 1.14–1.32), and participants in the highest DNN-IR quartile had a 77% increased risk compared with the lowest quartile (OR = 1.77, 95% CI: 1.43–2.20). These effect sizes are comparable to those reported for traditional IR indices^[30–32]^. For example, in the Bruneck Study, HOMA-IR was associated with an approximately 40% increased CVD risk per 1-SD increment after multivariable adjustment^[30]^. A recent study using the UK Biobank demonstrated that eGDR predicted CVD risk across stages 0–3 of the cardiovascular-kidney-metabolic syndrome^[32]^. The consistency of our findings with these reports supports the biological validity of DNN-IR as an IR assessment tool.

The dose-response relationship between DNN-IR and incident CVD, as assessed by restricted cubic spline regression, was approximately linear across the full range of DNN-IR values (*P*-overall < 0.001; *P*-nonlinear = 0.453), with no evidence of a threshold effect. This linear pattern was consistently observed for both incident heart disease and stroke. Notably, for heart disease, the risk remained stable at lower DNN-IR scores (< −1.0) but rose sharply once DNN-IR exceeded the median, suggesting a potential inflection point beyond which IR-mediated cardiovascular risk accelerates. This finding has clinical implications for risk stratification, as it identifies a subpopulation that may benefit most from targeted preventive interventions.

Importantly, the association between DNN-IR and incident CVD risk remained consistent across all prespecified subgroups except the education level stratum— encompassing age, sex, smoking and drinking status, SBP, DBP, TG, TC, HDL-C, and LDL-C—with no statistically significant interactions detected (all *P* for interaction > 0.05). This uniformity robustly supports the generalizability of DNN-IR as a CVD risk prediction tool, validating its utility across diverse population subgroups.

### DNN-IR and Mortality Outcomes

In the external validation cohort from the NHANES, the DNN-IR demonstrated robust predictive performance for mortality outcomes. The DNN-IR achieved an AUC of 0.77 for cardiovascular mortality and 0.72 for all-cause mortality, indicating substantial prognostic value that is consistent with previous NHANES-based analyses^[33,34]^. Furthermore, Kaplan-Meier analysis confirmed that the cumulative incidence of both CVD and all-cause mortality was significantly elevated in the highest DNN-IR quartile (log-rank *P* < 0.0001 for all), reinforcing the prognostic utility of the DNN-IR at the population level.

Of particular note, the DNN-IR exhibited outstanding predictive accuracy across a diverse spectrum of cause-specific mortalities, achieving remarkably high AUC values for diabetes-related (0.91), Alzheimer’s disease (0.88), chronic lower respiratory disease (0.86), and nephritis-related (0.96) mortality. While the mechanistic link between IR and diabetes-related mortality is well-established, its strong predictive capability for neurodegenerative and renal disease mortality is equally compelling. Accumulating evidence underscores the close association between insulin resistance and the pathogenesis of Alzheimer’s disease, mediated by accelerated cerebral amyloid-β deposition, tau protein hyperphosphorylation, exacerbated neuroinflammation, and impaired cerebral glucose metabolism^[35–37]^.

Moreover, the exceptional predictive performance of the DNN-IR for nephritis-related mortality aligns with the established role of IR in renal dysfunction and the emerging paradigm of "cardiovascular-kidney-metabolic (CKM) syndrome" ^[32,38]^. The ability of a single IR index to predict mortality risk across such a wide array of distinct disease categories suggests that, compared with traditional IR indices, the DNN-IR more comprehensively captures the systemic and multi-organ pathophysiological consequences driven by insulin resistance.

### Biological Plausibility and the MoE Architecture

The biological plausibility of the DNN-IR components is well-established. FPG and HbA1c reflect glycemic dysregulation, the core of IR. Body weight and WC capture adipose-driven IR, where central obesity induces lipotoxicity and inflammation that impair peripheral insulin signaling^[39]^. Hypertension exists in a bidirectional cycle with IR; hyperinsulinemia exacerbates vascular stiffness via sodium retention and sympathetic activation^[40]^.

Integrating these components via a Mixture of Experts (MoE) architecture advances beyond traditional fixed-formula indices (e.g., HOMA-IR, TyG). The MoE framework dynamically weights distinct subnetworks via a gating mechanism to capture specific IR phenotypes^[41]^. This is ideal for modeling the heterogeneity of IR, as the relative contributions of metabolic and vascular factors vary across subpopulations. Additionally, the sparse mask mechanism filters the most predictive features, yielding a parsimonious, highly interpretable five-variable index with strong clinical utility.

### Comparison with Previous Machine Learning-Based IR Approaches

Several studies have explored ML and deep learning (DL) approaches for IR assessment^[42–44]^. Tsai et al. developed an ML-based IR prediction model for non-diabetic populations and demonstrated its association with incident CVD and mortality^[43]^. Peng et al. used LightGBM to predict IR in non-diabetic populations and validated its clinical value in cohort settings^[44]^. More recently, Metwally et al. demonstrated that IR could be predicted from wearable device data and routine blood biomarkers using deep learning^[42]^.

Our study extends these findings in several important ways. First, we incorporated baPWV — a direct measure of arterial stiffness — as the target label, rather than relying on HOMA-IR or clamp-derived measures as surrogate labels, which may introduce circularity when the downstream outcome of interest is CVD. Second, the MoE architecture with sparse masking represents a methodological innovation that simultaneously optimizes predictive performance and variable selection. Third, we conducted external validation in two geographically and ethnically distinct cohorts (CHARLS in China, NHANES in the US), providing evidence of cross-population generalizability. Fourth, our comprehensive evaluation of 19 existing IR indices alongside DNN-IR establishes a clear benchmark for future IR index development.

### Strengths and Limitations

This study has several notable strengths. First, the utilization of three large, well-characterized cohorts—REACTION (derivation), CHARLS (external validation for incident CVD), and NHANES (external validation for mortality)—provides robust, multi-source evidence. The REACTION study offered a large-scale Chinese population sample with comprehensive baPWV measurements, enabling the development of an atherosclerosis-targeted IR index. The inclusion of 19 traditional IR indices as comparators allowed for rigorous benchmarking. The integration of a sparse mask mechanism with a MoE deep learning framework represents a methodological advancement over conventional regression-based approaches. Furthermore, sensitivity analyses confirmed the robustness of our primary findings against multicollinearity, and subgroup analyses demonstrated the consistency of the associations across diverse population strata.

Several limitations should be acknowledged. First, the derivation cohort (the REACTION Fujian subgroup) was restricted to a single geographic region in China, which may limit generalizability to other ethnic populations, although external validation in CHARLS and NHANES partially mitigates this concern. Second, while baPWV is a well-validated surrogate marker for arterial stiffness, it is not equivalent to vascular angiography, the gold standard for assessing structural atherosclerosis. However, we intentionally selected baPWV as the target label because it non-invasively and directly reflects the functional vascular consequences of IR that are most relevant to CVD prediction. Third, the deep learning model was trained on cross-sectional data from the REACTION study; future research should explore longitudinal IR trajectory modeling. Fourth, differences in baseline characteristics, data collection protocols, and follow-up durations among the CHARLS and NHANES cohorts may affect cross-cohort comparability. Fifth, despite comprehensive adjustment for confounders, residual confounding from unmeasured variables (e.g., diet, physical activity, and socioeconomic factors) cannot be entirely excluded. Sixth, the calculation of DNN-IR requires five clinical variables; although routinely measured in clinical practice, its convenience may be inferior to single-biomarker indices such as TyG in resource-limited settings.

### Conclusions

In this study, we developed DNN-IR, a novel deep learning-derived insulin resistance index that accurately identifies atherosclerosis using only five routine clinical variables, outperforming 19 traditional indices. Across multiple external cohorts, DNN-IR robustly predicted incident CVD and various cause-specific mortalities, exhibiting a linear dose-response relationship. As a comprehensive biomarker of metabolic health, DNN-IR provides a highly practical and novel tool for the early risk stratification of cardiometabolic diseases in clinical practice.

## Data Availability

Portions of the data supporting this study are publicly available from the National Health and Nutrition Examination Survey (NHANES) website (https://www.nhanescdc.gov/nchs/nhanes/) and the China Health and Retirement Longitudinal Study (CHARLS) website (https://charls.pku.edu.cn/). Data from the Fujian subcohort of the REACTION (Risk Evaluation of Cancers in Chinese Diabetic Individuals: A Longitudinal Study) are available from the corresponding author upon reasonable request.

## Acknowledgements

This study utilized data from the China Health and Retirement Longitudinal Study (CHARLS), the National Health and Nutrition Examination Survey (NHANES), and the Fujian sub-cohort of the Risk Evaluation of cAncers in Chinese diabeTic Individuals: a lONgitudinal study (REACTION). The authors extend sincere gratitude to the staff members of the CHARLS, NHANES, and REACTION project teams, as well as all participants involved in these surveys.

## Author contributions

YQM and JL performed data analysis and drafted the manuscript; YQM, AZ, SHZ, and DY conducted data cleaning; WJY constructed the deep learning models and performed model analyses. YQM, WL, JPW, and GC designed the study, JPW, and GC provided overall supervision. All authors actively participated in the research process, made substantial contributions to manuscript revision, and carefully reviewed and approved the final version.

## Ethics approval and consent to participate

The REACTION study protocol was approved by the Ethics Committee of Fuzhou University Affiliated Provincial Hospital. Written informed consent was obtained from all participants. Regarding the NHANES and CHARLS databases, this study exclusively utilized anonymized public datasets that do not involve direct intervention or individual identification. In accordance with relevant ethical guidelines, obtaining additional informed consent was not required. The researchers strictly adhered to the NHANES and CHARLS Data Use Agreements to ensure compliance with ethical standards and data security protocols.

